# The correspondence between the structure of the terrestrial mobility network and the emergence of COVID-19 in Brazil

**DOI:** 10.1101/2020.05.17.20104612

**Authors:** Vander L. S. Freitas, Thais C. R. O. Konstantyner, Jeferson Feitosa, Catia S. N. Sepetauskas, Leonardo B. L. Santos

## Abstract

**BACKGROUND:** the inter-cities mobility network serves as a proxy for the SARS-CoV-2 spreading network in a country.

**OBJECTIVE:** to investigate the correspondences between the structure of the mobility network and the emergence of COVID-19 cases in Brazilian cities.

**METHODS:** we adopt the data from the Brazilian Health Ministry and the terrestrial flow of people between cities from the IBGE database in two scales: Brazilian cities without the North region and cities from the Sao Paulo state. Grounded on the complex networks approach, cities are represented as nodes and the flows as edges. Network centrality measures such as strength and degree are ranked and compared to the list of Brazilian cities, ordered according to the day that they confirmed the first case of COVID-19.

**FINDINGS:** The strength presents the best correspondences and the interiorization process of SARS-CoV-2 is captured in the Sao Paulo state when different thresholds are applied to the network flows.

**MAIN CONCLUSIONS:** the strength captures the cities with a higher vulnerability of receiving new cases of COVID-19. Some countryside cities such as Feira de Santana (Bahia state), Ribeirao Preto (Sao Paulo state), and Caruaru (Pernambuco state) have strength comparable to states’ capitals, making them potential super spreaders.

**Financial support:** São Paulo Research Foundation (FAPESP) Grant Numbers 2015/50122-0, 2018/06205-7 and 2020/06837-3; DFG-IRTG Grant Number 1740/2; CNPq Grant Numbers 420338/2018-7 and 101720/2020-3.

## INTRODUCTION

The world is currently facing a global public health emergency due to the COVID-19 pandemic, declared on March 11th, 2020 by the World Health Organization (WHO). The outbreak began in late December 2019 in Wuhan, Hubei province in China, spreading rapidly to several countries^(1)^.

As of June 4th, 2020, more than 6.7 million cases have been confirmed worldwide, with almost 400 thousand deaths. In Brazil, the first documented case was in the city of São Paulo on February 26th, 2020. Since then, there are about 615,000 confirmed cases and 34,000 deaths in the national territory^(2,3,4)^.

The inter-cities mobility network serves as a proxy for the transmission network, vital for understanding outbreaks, especially in Brazil, a continental-dimension country^(5,6,7,8,9)^. The complex network approach^(10)^ emerges as a natural mechanism to handle mobility data computationally, taking areas as nodes (fixed) and movements between origins and destinations as connections (flows). Some networks’ measures can be used to find the structurally more vulnerable areas in the context of the current study. The degree of a node is the number of cities that it is connected to, showing the number of possible destinations. The strength captures the total number of people that travel to (or come from) such places in a given time frame. From a probability perspective, the cities that receive more people are more vulnerable to SARS-CoV-2. The betweenness centrality, on the other hand, considers the entire network to depict the topological importance of a city in the routes that are more likely to be used.

The present work aims to investigate the correspondences of the networks’ measures with the emergence of cities with confirmed cases of COVID-19 in two scales: Brazil and the state of São Paulo. Specifically, we analyze i) the Brazilian inter-cities mobility networks under different flow thresholds to neglect the lowest-frequency travels, especially in the beginning of the outbreak, when the interiorization of the disease is still not in progress; and ii) the correspondence between the networks’ statistics and the emergence of COVID-19 in Brazil.

## METHOD

The most common mobility data used in studies of this nature in Brazil are the pendular travels, from the 2010 national census (IBGE)^(11)^. In this paper, we use the roads’ IBGE data from 2016^(12)^, which contains the flows between cities considering terrestrial vehicles in which it is possible to buy a ticket (mainly buses and vans). The information collected by that research seeks to quantify the interconnection between cities, the movement of attraction that urban centers carry out for the consumption of goods and services, and the long-distance connectivity of Brazilian cities. The North region is not included in this paper, because neither the fluvial nor the air modals are covered, and their roles are crucial to understanding the spreading process there, especially in the Amazon region.

The above-cited IBGE data^(12)^ contains the weekly travel frequency (flow) between pairs of Brazilian cities/districts. The frequencies are aggregated within the round trip, which means that the number of travels from city A to city B is the same as from B to A. We produce two types of undirected networks with a different number *N* of nodes to capture actions in two scales (country and state):

1. *N* = 4987 - Brazil without the North region (BRWN): nodes are cities and edges are the flow of direct travels between them.
2. *N* = 620 - São Paulo state (SP): a subset of the previous network, containing only cities within the São Paulo state.

Some cities are not present in our networks, due to a simplification that the IBGE does: it groups small neighboring municipalities with almost no flow into single nodes. For simplicity, and considering that such places do not contain cases in the first days of the outbreak, they are not individually accounted for in our analysis.

We focus on two versions of each network for certain flow thresholds η, the η_0_ (η = 0) that is the original network from the IBGE data and η_*d*_ (η = *d*), to neglect travels with lower-level frequencies. The *d* corresponds to the higher flow threshold that produces the network with the largest diameter. The motivation behind η_*d*_ is to get a threshold high enough to not consider the least frequent connections and to not disregard the most frequent ones^(8,13)^.

According to the notified cases from daily state bulletins of the Brazilian Health Ministry^(4)^, until June 4th, 2020, the number of cities with at least one confirmed patient with COVID-19 is *X* = 3851 in the BRWN network, which corresponds to 77% of the nodes, and *X* = 535 in SP (86% of the nodes). We use this data to track the response of each measure in detecting vulnerable cities according to the evolution of the virus spreading process.

### Complex network measures

The topological degree^(10)^ *k* of a node is the number of links it has to other nodes. As here the networks are undirected, there is no distinction between incoming and outgoing edges.

In a connected graph, there is at least one shortest path σ_*vw*_ between any pair of nodes *v* and *w*. The betweenness centrality^(10)^ *b* of a node *i* is the rate of those shortest paths that pass through *i*:

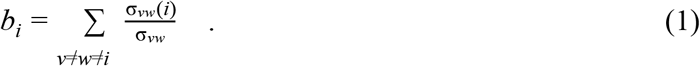

Although it is a pointwise measure, it takes into account non-local information related to all shortest paths on the network. It is worth highlighting that in the present context this centrality index is not a transportation (physical) measure but a mobility (process) one. Besides, both degree and betweenness do not account for the network flows here, but the binary (weightless) networks. The diameter of a network is the distance between the farthest nodes, given by the maximum shortest path.

The strength^(10)^ of a node on the other hand is the accumulated flow from incident edges:

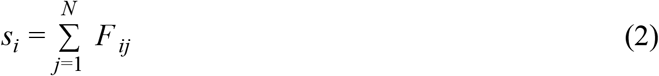

in which *F* _*ij*_ is the flow between nodes *i* and *j*.

Figure 1 presents illustrations of very simple networks with the aforementioned measures calculated for each node. The bigger and more red the nodes, the higher the values associated with them. In Figure 1a, node F has the highest strength, meaning that it receives more flow than any other. Nodes B, D and E are the most connected, each with exactly four incident edges - see Figure 1b. Lastly, node F of Figure 1c has the higher betweenness value, since all shortest paths with end on G pass through it. Nodes D and E both divide the very same load of shortest paths and have intermediate values.

**Figure 1:**
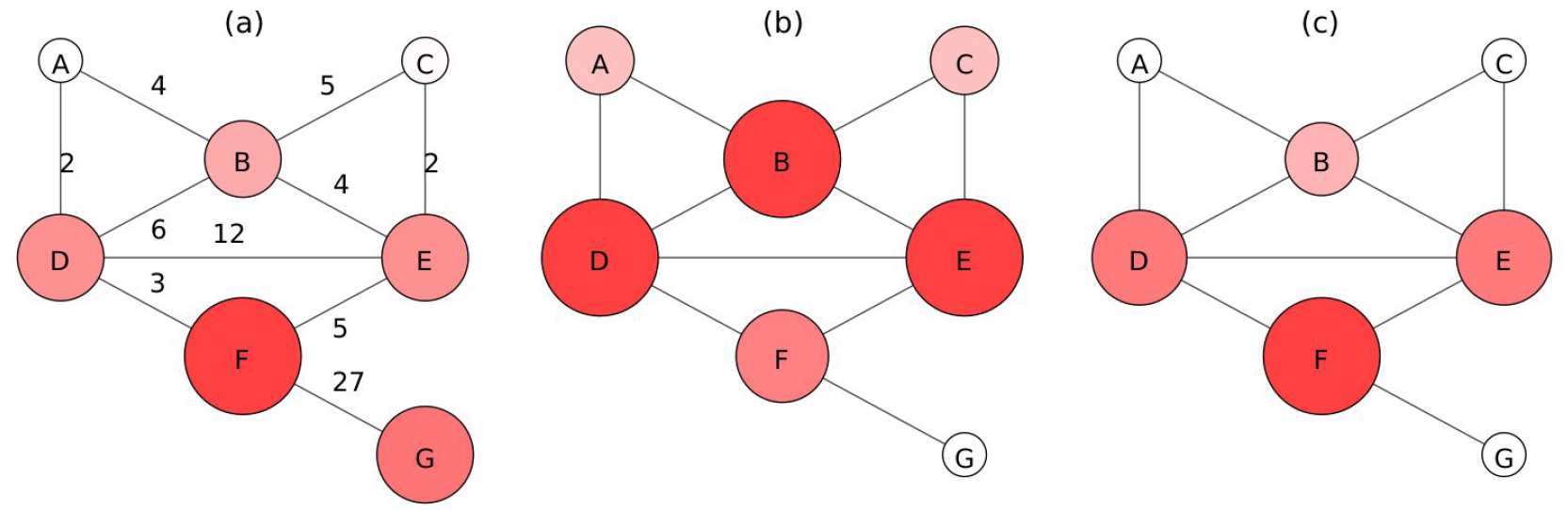
Illustration of network measures: a) strength, b) degree, and c) betweenness. Size and colors are related to the node measure. The network of subfigure *a* is weighted, while the others not.

### Geographical visualization

A geographical approach for complex systems analysis is especially important for mobility phenomena. Santos *et al*. (2017)^(14)^ proposed a graph where the nodes have a known geographical location, and the edges have spatial dependence, the (geo)graph. It provides a simple tool to manage, represent, and analyze geographical complex networks in different domains^(8,13)^ and it is used in the present work. The geographical manipulation is performed with the PostgreSQL Database Management System and its spatial extension PostGIS. Lastly, the maps are produced using the Geographical Information System ArcGIS.

## RESULTS

This section presents the results of the topological analysis for the previously mentioned networks. Table 1 showcases the size *N* of each network, number of edges |*E*|, average strength ⟨*s*⟩, average degree ⟨*k*⟩, and average betweenness ⟨*b*⟩.

**TABLE 1.**
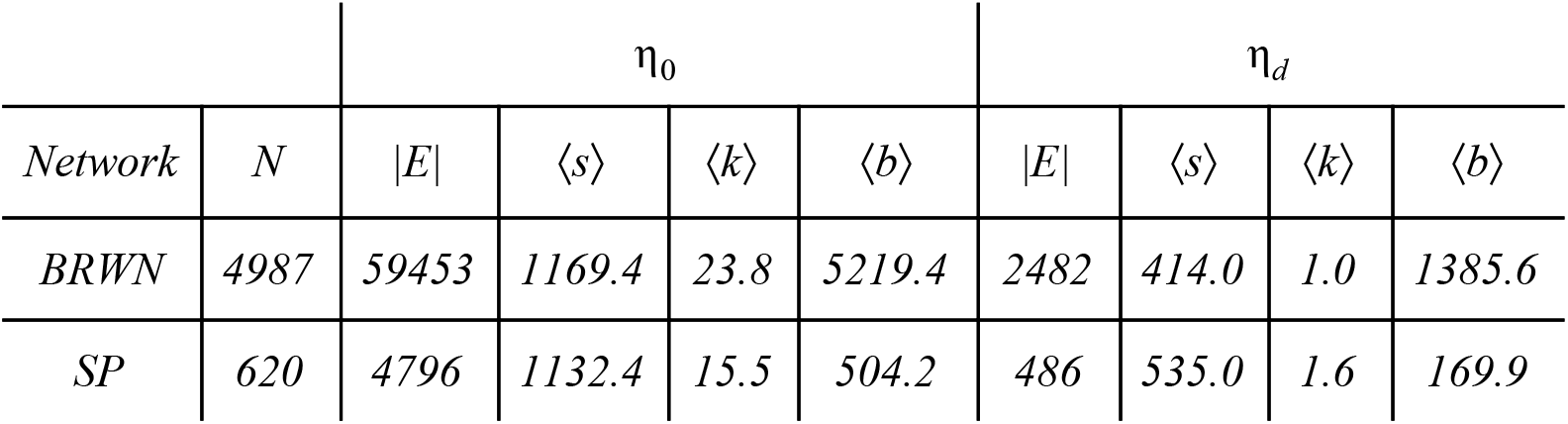
Statistics for the Brazilian (BRWN) and São Paulo state (SP) networks, with two flow thresholds: η_0_ (original flows) and η_d_ (the higher threshold with maximum diameter). For each network and threshold, the number of edges |E|, number of nodes N, average degree ⟨k⟩, average strength ⟨s⟩, and average betweenness ⟨b⟩ are displayed.

| Network | $N$ | $\eta_0$ | | | | $\eta_d$ | | | |
| --- | --- | --- | --- | --- | --- | --- | --- | --- | --- |
| | | $\vert E\vert $ | $\langle s \rangle$ | $\langle k \rangle$ | $\langle b \rangle$ | $\vert E\vert $ | $\langle s \rangle$ | $\langle k \rangle$ | $\langle b \rangle$ |
| BRWN | 4987 | 59453 | 1169.4 | 23.8 | 5219.4 | 2482 | 414.0 | 1.0 | 1385.6 |
| SP | 620 | 4796 | 1132.4 | 15.5 | 504.2 | 486 | 535.0 | 1.6 | 169.9 |

Two nodes are connected when between them there is a nonzero flow, which means that the number of connections |*E*| decreases for increasing threshold (η). The resulting networks are undirected and, throughout the paper, both the degree and the betweenness measures do not account for the flows, but weightless edges instead. The diameter of the networks for varying η is computed and Figure 2 shows the exact point (dashed line) where the higher threshold with maximum diameter is found for both networks: η_*d*_ = 207.55 for BRWN and η_*d*_ = 161.01 for SP.

**Figure 2:**
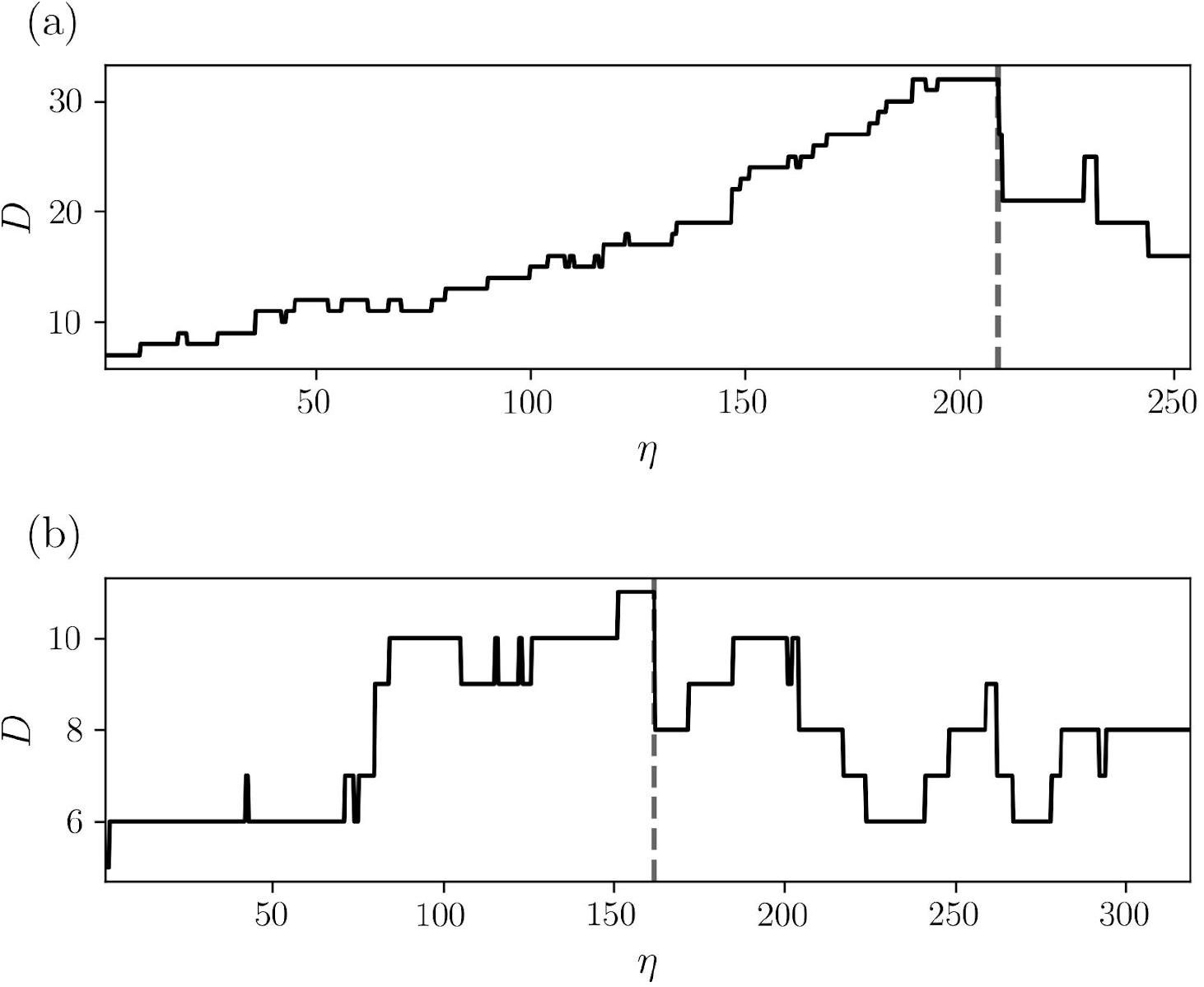
Diameter of the networks as a function of flow threshold η. The dashed line is the maximum diameter with higher η, with a) η_*d*_ = 207.55 for BRWN, and b) η_*d*_ = 161.01 for SP.

Following the (geo)graphs approach, it is possible to visualize nodes and edges of the Brazilian mobility network in the geographical space for η_*d*_ in Figure 3. The edges for η_0_ are not plotted, because there are more than 59000 and the visualization is not clear. It is important to highlight some key cities like Belo Horizonte, Rio de Janeiro, São Paulo and Salvador, and the high number of connections between them. Figure 4 depicts the geographical graph regarding the state of São Paulo.

**Figure 3:**
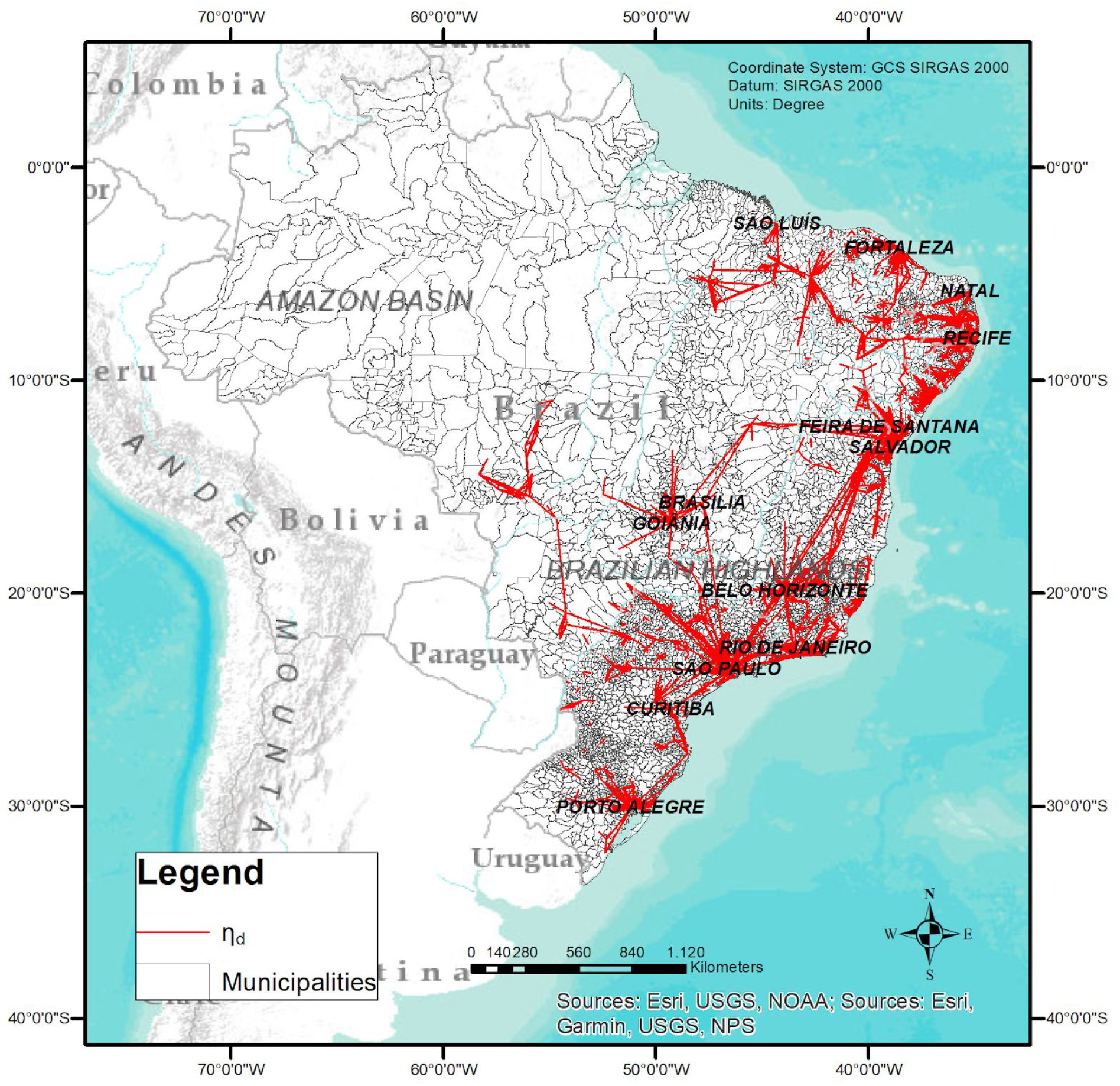
Map of the Brazilian municipalities with the sets of edges for η_*d*_. The edges for η_0_ are not plotted, because there are more than 59000 and the visualization was not clear.

**Figure 4:**
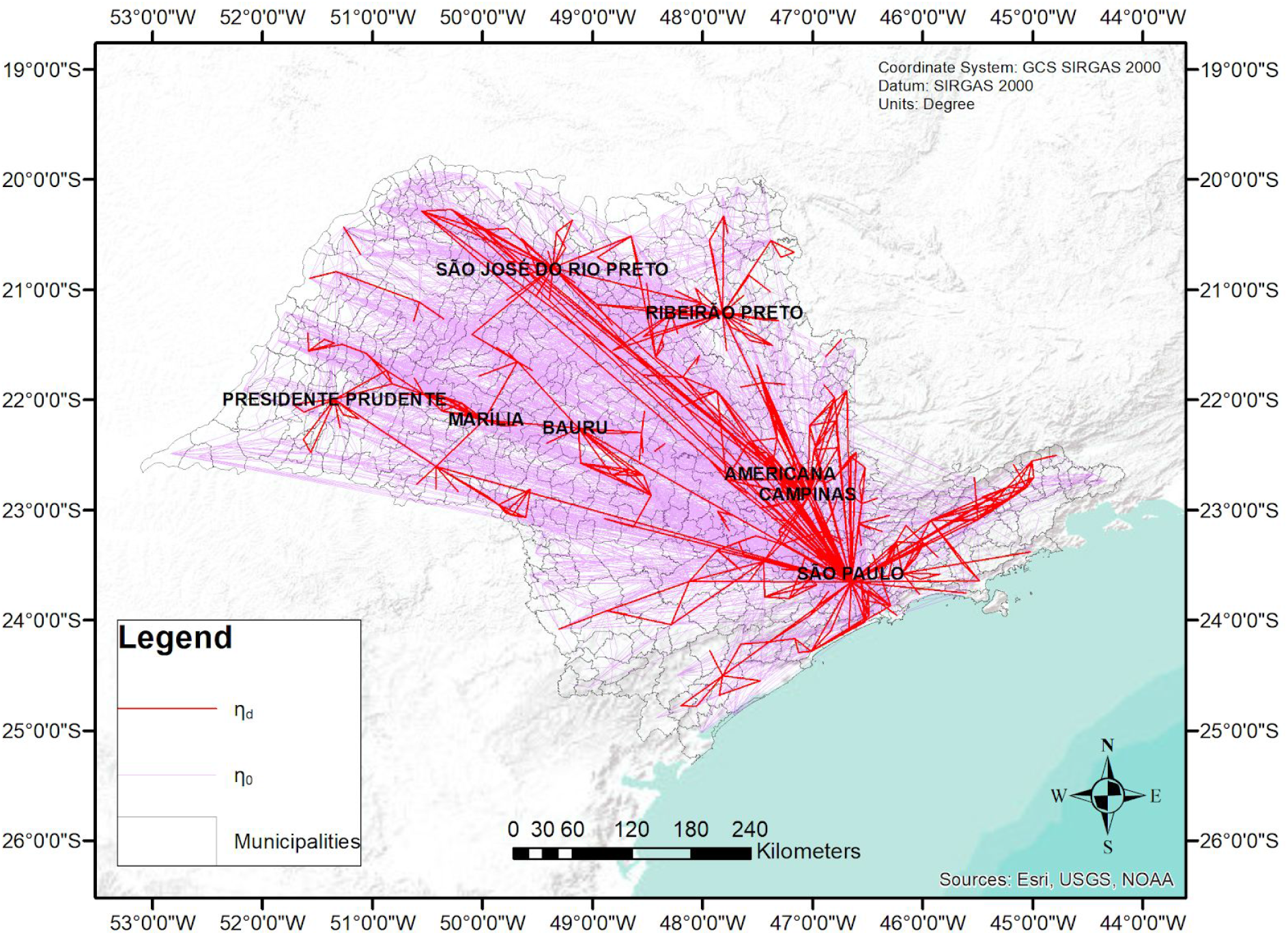
Map of the municipalities of the São Paulo state with the sets of edges for η_0_ and η_*d*_.

Figure 5 shows the map of the topological degree related to each node/city, considering all original flows (η_0_), and in Figure 6 there is the equivalent for η_*d*_ = 207.55. Key cities are labeled in the maps.

**Figure 5:**
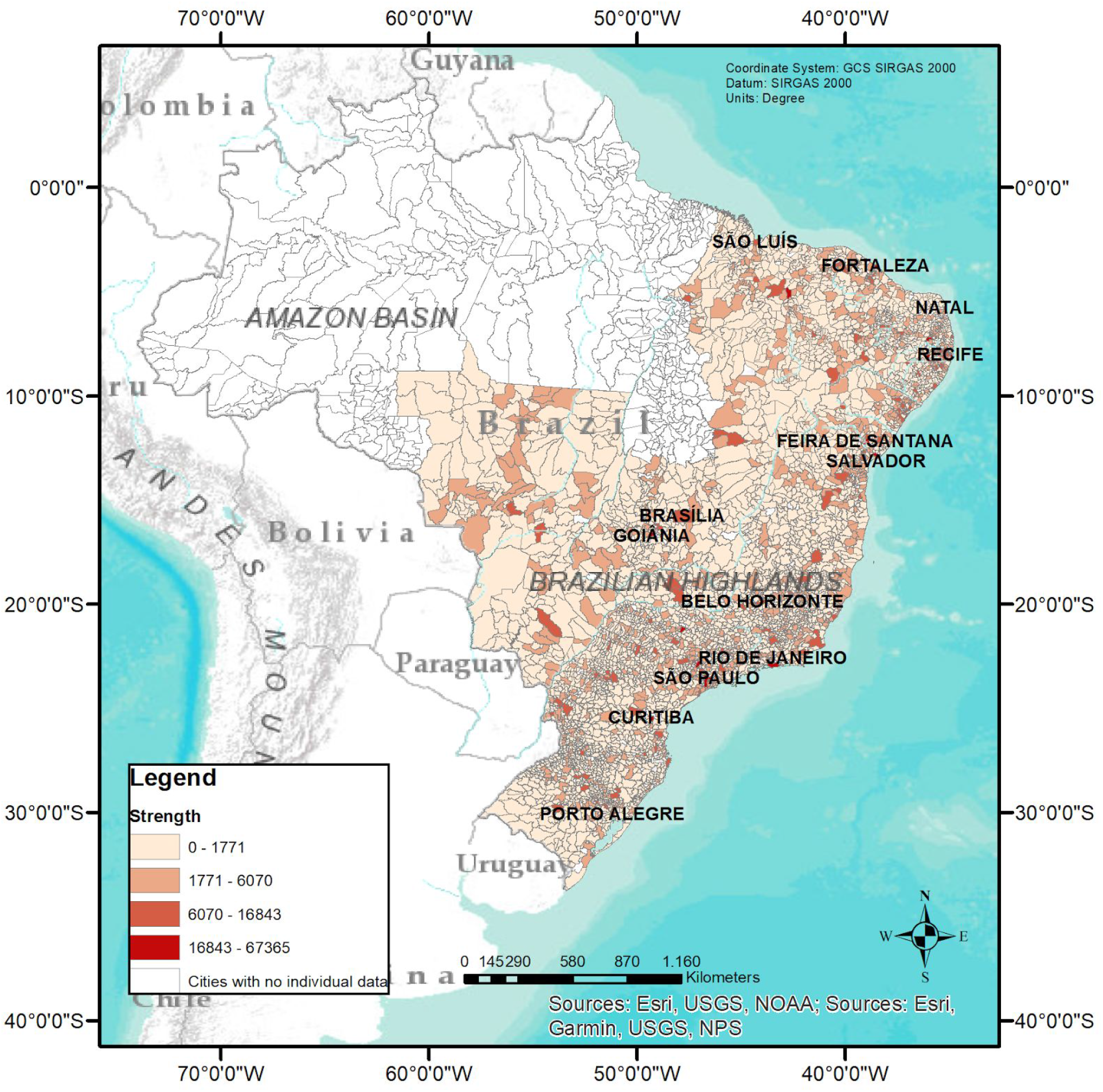
Map of the topological strength related to each node/city of the BRWN network, for η_0_.

**Figure 6:**
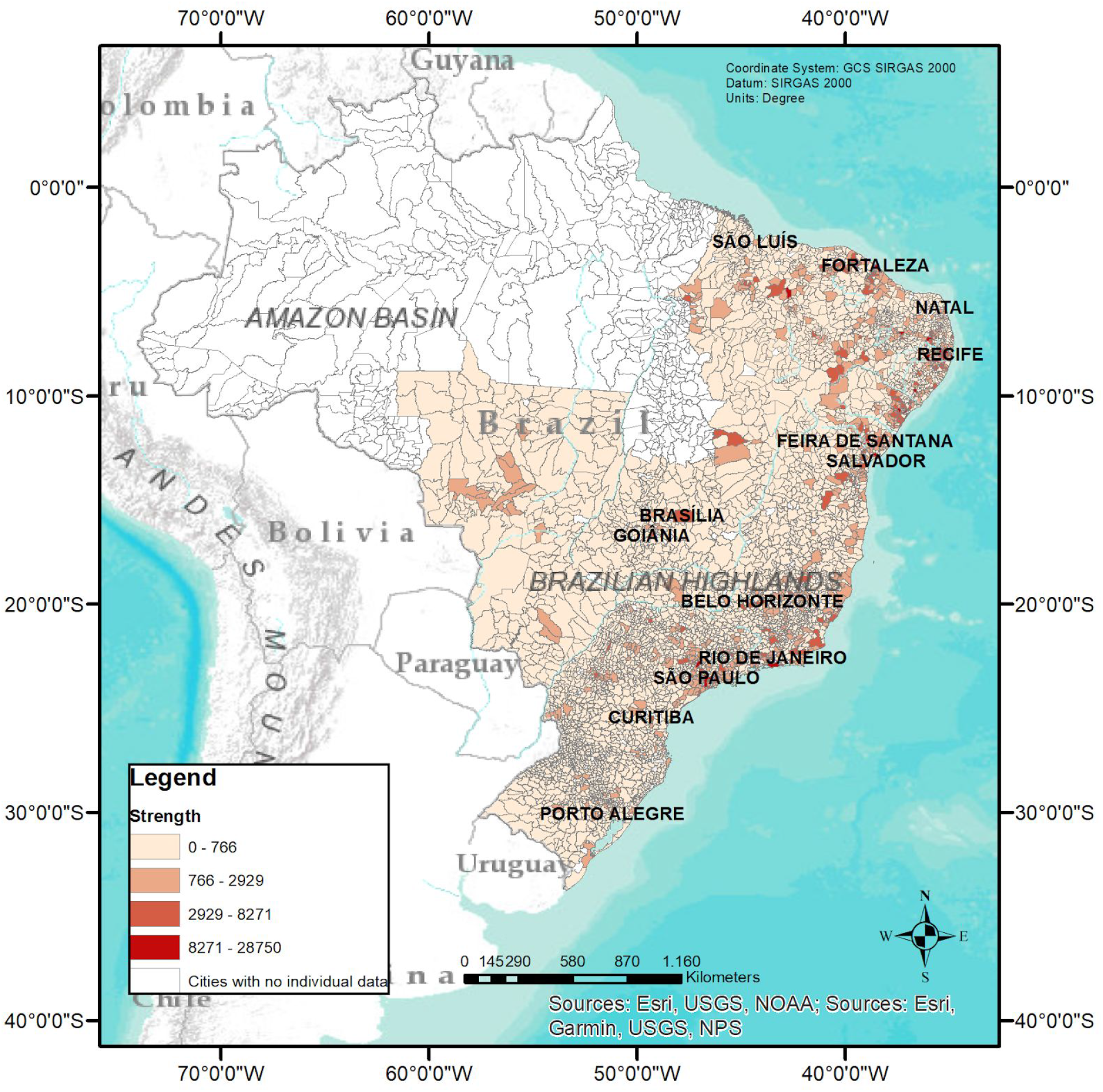
Map of the topological strength related to each node/city of the BRWN network, for η_*d*_.

Figures 7 and 8 present the strength for the SP network, with η_0_ and η_*d*_, respectively. Some cities with high strength also appear in a report^(17)^ of most vulnerable cities to COVID-19 due to their intense traffic of people, namely São Paulo, Campinas, São José do Rio Preto, São José dos Campos, Ribeirão Preto, Santos, Sorocaba, Jaboticabal, Bragança Paulista, Presidente Prudente, Bauru, and many others. Currently, they all have a significant number of confirmed cases.

**Figure 7:**
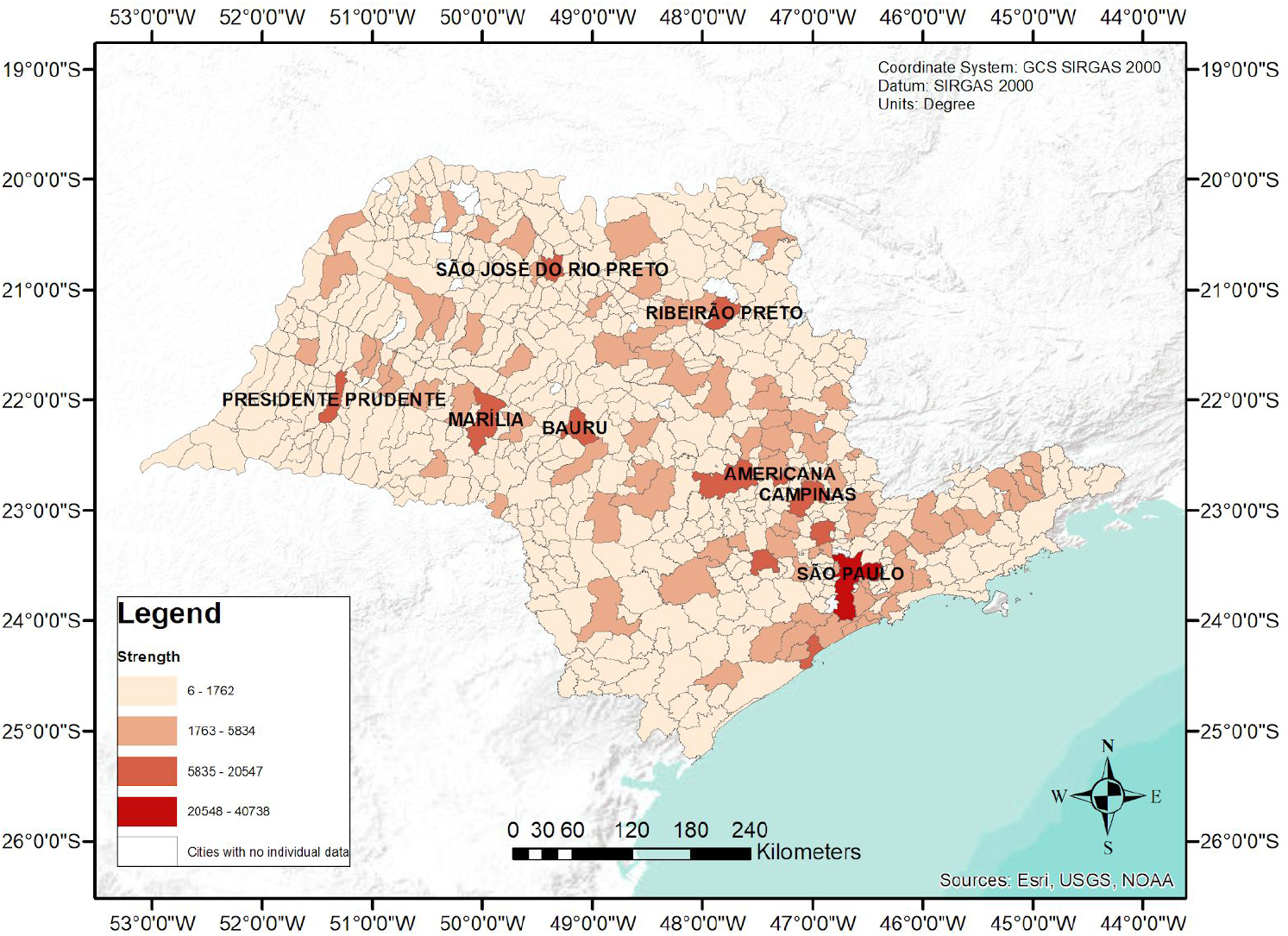
Map of the topological strength related to each node/city of the SP network, for η_0_.

**Figure 8:**
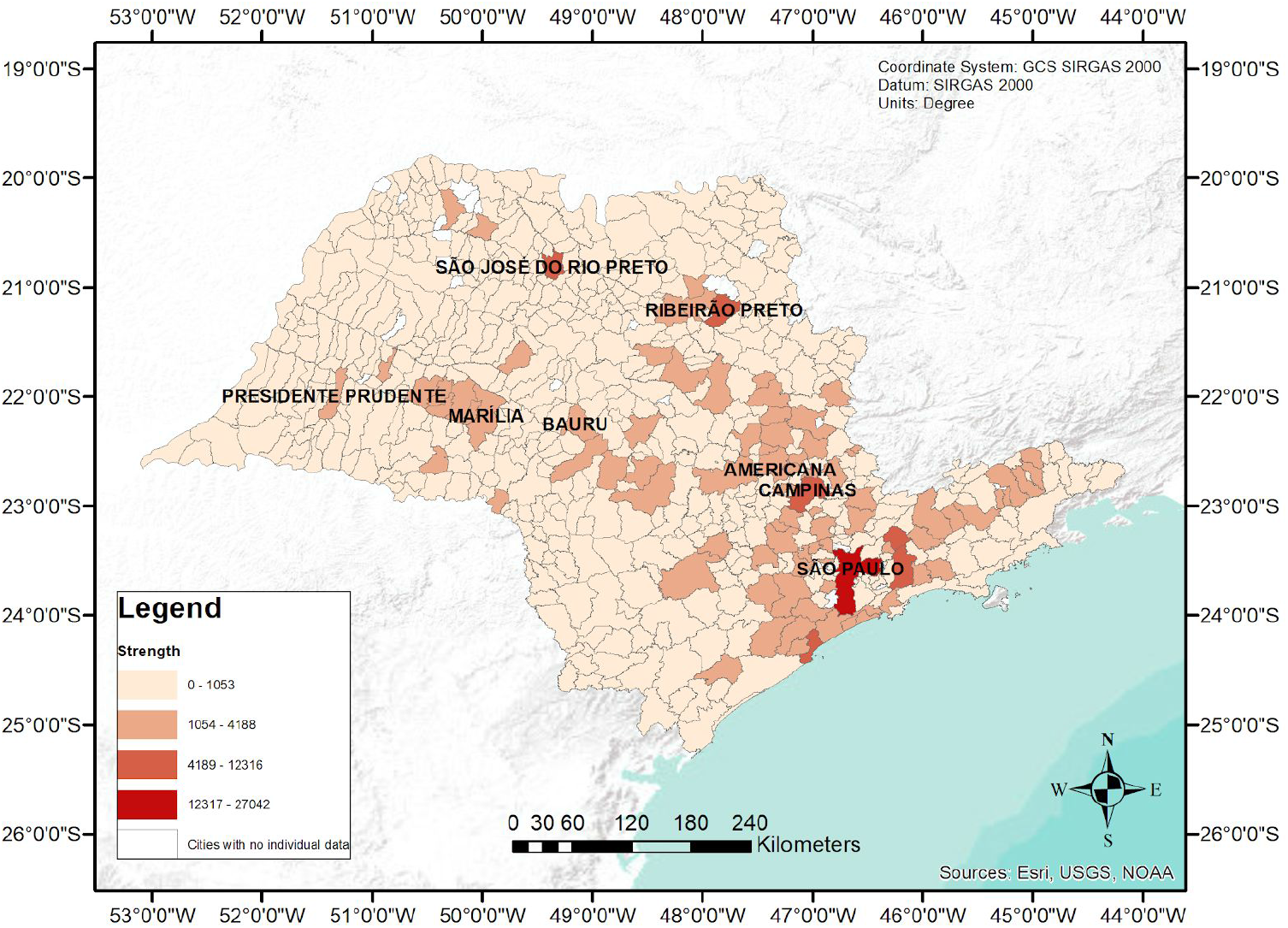
Map of the topological strength related to each node/city of the SP network, for η_*d*_.

We now assess which of the computed measures (*s, k*, and *b*) better approximates the emergence of COVID-19 in Brazil. We compare the top-ranked *n* ∈ [1, *X*] cities of each measure with the *n* cities that contain confirmed cases. The available data of the notified cases from daily state bulletins of the Brazilian Health Ministry^(4)^, until june 4th, 2020, presents *X* = 3851 in the BRWN network, which corresponds to 77% of the nodes, and *X* = 535 in SP (86% of the nodes).

In order to verify whether the rate of correspondence between the top-ranked cities from the networks’ measures and the cities with COVID-19 cases has statistical significance, we verify what are the results of picking cities at random instead of under the measures’ guidance. We perform 10^5^ simulations for each *n* ∈ [1, *X*], choosing *n* nodes by sort and monitoring what is the rate *p* of positive cases. Figure 9 presents the correspondence of the first *n* cities with COVID-19 documented cases and both the simulated data and the top-ranked nodes under *s, k*, and *b*. The gray region represents 95% of the rates’ occurrences in the simulations for each *n*.

**Figure 9:**
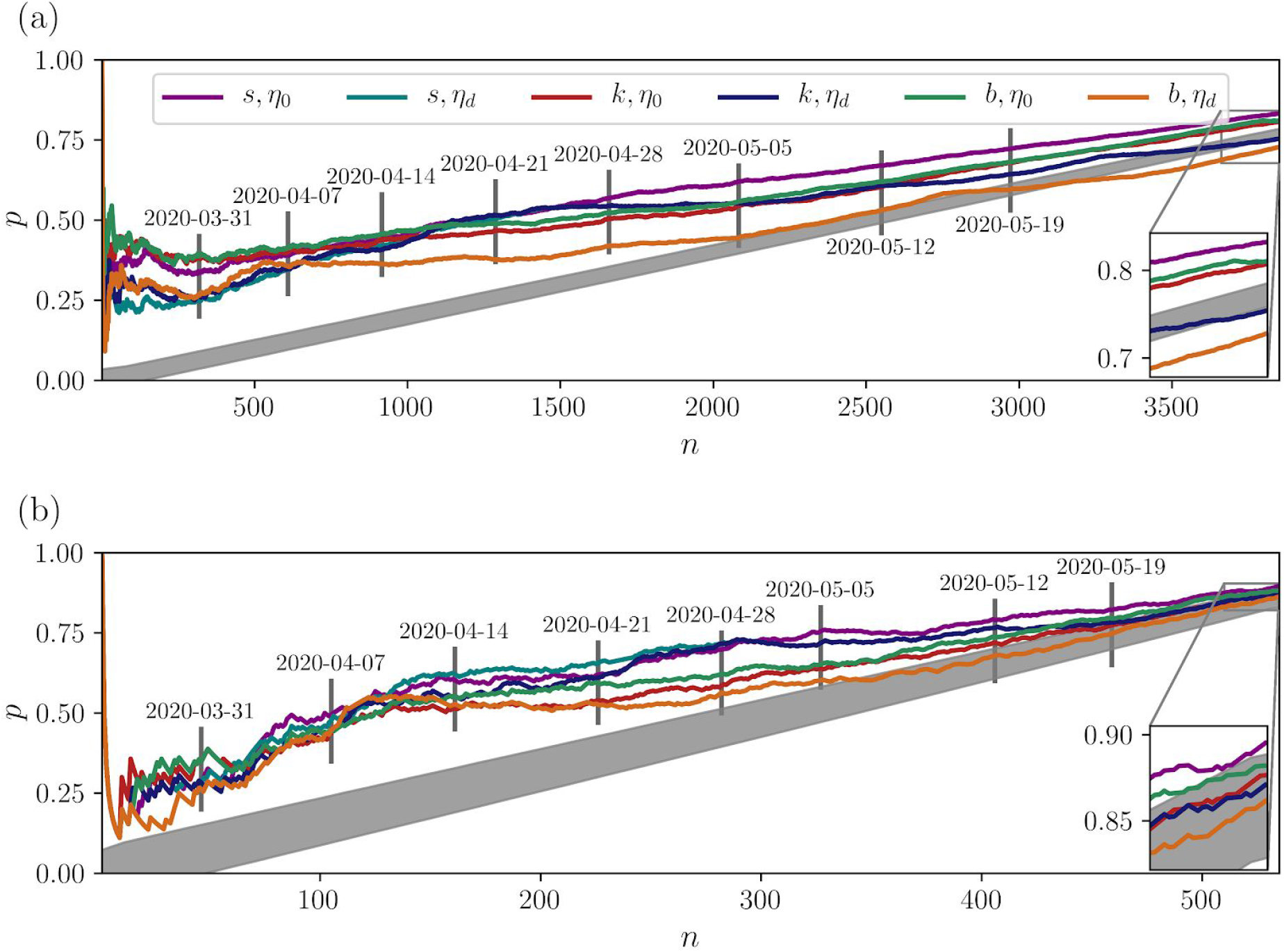
Correspondence (rate) between the *n* top ranked cities for different network criteria: *s, k*, and *b*, and cities that have at least one patient with COVID-19 in: a) BRWN and b) SP. The gray region represents the correspondences with randomly selected cities. The inset is a zoomed area of the last days until June 4th. In both panels, the *s* and *k* with η_*d*_ are overlapping in the end.

In our analysis, on June 4th, about 95% of the simulations have matching rates within 0.77 ± 0.01 for the BRWN network, and the same volume is within 0.86 ± 0.03 for the SP. The results for node selection during the first days via the network indexes all lie above the gray region, which means that all indexes are a better heuristic than picking nodes at random in the beginning. However, immediately after May 5th, *b* with η_*d*_ starts to touch the region in SP, having, therefore, results compared to the simulations. It has the worst results for BRWN as well, after a transient.

High-frequency oscillations are perceived in Figure 9 for small *n*, but they stabilize afterward and follow a tendency. In Figure 9a they are pronounced until March 24th (*n* ≈ 150). The matching *p* is at maximum in the beginning, because the first documented case was in the city of São Paulo, which is the first ranked city in all measures. The curve then decreases until reaching a region where the oscillations take place. Furthermore, the network quantifiers pose good correspondences already in the beginning of the spreading process as the gray regions is not touched until *n* approaches *X*.

Interestingly, on March 31st, the high-frequency oscillations start to diminish in SP. A few days further, after April 7th, the betweenness centrality with η_*d*_ starts to be a bad predictor for BRWN and then for SP.

Table 2 enumerates the first twenty ordered cities according to the best-evaluated measures and compares them side-by-side with the first twenty cities with COVID-19 cases in the BRWN network. The best measures for SP are compared with each other in Table 3 as well. Although in this case *s* with η_0_ presents good correspondences, we present the ones with η_*d*_ due to its importance until the end of April. In both networks, although *p* produces high-frequency oscillations in the beginning, as shown in Figure 9, the metrics still pose some correspondences with the first confirmed cases.

**TABLE 2.** Cities with at least one case of COVID-19 in Brazil (BRWN) in the order they were documented^(10)^, side-by-side with the top-ranked cities regarding s, k and b for η_0_ and η_d_. The best combination is s with η_0_ (second column). Matching cities are colored alike.

| | COVID-19 | $s, \eta_0$ | $k, \eta_0$ | $b, \eta_0$ |
| --- | --- | --- | --- | --- |
| 1 | 3550308 São Paulo | 3550308 São Paulo | 3550308 São Paulo | 3550308 São Paulo |
| 2 | 3300407 Barra Mansa | 3106200 Belo Horizonte | 3106200 Belo Horizonte | 3106200 Belo Horizonte |
| 3 | 2910800 Feira de Santana | 2927408 Salvador | 3509502 Campinas | 5208707 Goiânia |
| 4 | 3304557 Rio de Janeiro | 2800308 Aracaju | 5300108 Brasília | 5300108 Brasília |
| 5 | 5300108 Brasília | 3509502 Campinas | 5208707 Goiânia | 3304557 Rio de Janeiro |
| 6 | 3547304 Santana de Parnaíba | 2304400 Fortaleza | 3304557 Rio de Janeiro | 4314902 Porto Alegre |
| 7 | 3122306 Divinópolis | 4314902 Porto Alegre | 4314902 Porto Alegre | 2304400 Fortaleza |
| 8 | 2704302 Maceió | 2504009 Campina Grande | 2927408 Salvador | 4106902 Curitiba |
| 9 | 4303905 Campo Bom | 2910800 Feira de Santana | 2910800 Feira de Santana | 3509502 Campinas |
| 10 | 4314902 Porto Alegre | 2611606 Recife | 4106902 Curitiba | 2408102 Natal |
| 11 | 4305108 Caxias do Sul | 2604106 Caruaru | 4104808 Cascavel | 2910800 Feira de Santana |
| 12 | 4105508 Cianorte | 5208707 Goiânia | 3543402 Ribeirão Preto | 2927408 Salvador |
| 13 | 4106902 Curitiba | 3304557 Rio de Janeiro | 2304400 Fortaleza | 2211001 Teresina |
| 14 | 3515707 Ferraz de Vasconcelos | 2604007 Carpina | 4115200 Maringá | 3543402 Ribeirão Preto |
| 15 | 4205407 Florianópolis | 2211001 Teresina | 4304705 Carazinho | 2111300 São Luís |
| 16 | 5208707 Goiânia | 3543402 Ribeirão Preto | 5201108 Anápolis | 2504009 Campina Grande |
| 17 | 3131307 Ipatinga | 4106902 Curitiba | 3549805 São José do Rio Preto | 2611606 Recife |
| 18 | 2408102 Natal | 2608909 Limoeiro | 2933307 Vitória da Conquista | 2507507 João Pessoa |
| 19 | 3303302 Niterói | 2610608 Paudalho | 2903201 Barreiras | 2800308 Aracaju |
| 20 | 2611606 Recife | 2510808 Patos | 5002704 Campo Grande | 3549805 São José do Rio Preto |

**TABLE 3.** Cities with at least one case of COVID-19 in the State of São Paulo (SP) in the order they were documented^(10)^, side-by-side with the top-ranked cities regarding s, k and b between η_0_ and η_d_. The best combination is s with η_d_ (second column) until the end of April. Matching cities are colored alike.

| | COVID-19 | $s, \eta_d$ | $k, \eta_d$ | $b, \eta_0$ |
| --- | --- | --- | --- | --- |
| 1 | 3550308 São Paulo | 3550308 São Paulo | 3550308 São Paulo | 3550308 São Paulo |
| 2 | 3547304 Santana de Parnaíba | 3509502 Campinas | 3509502 Campinas | 3509502 Campinas |
| 3 | 3515707 Ferraz de Vasconcelos | 3543402 Ribeirão Preto | 3543402 Ribeirão Preto | 3549805 S. José do Rio Preto |
| 4 | 3529401 Mauá | 3537602 Peruíbe | 3549805 S. José do Rio Preto | 3543402 Ribeirão Preto |
| 5 | 3547809 Santo André | 3549805 S. José do Rio Preto | 3552205 Sorocaba | 3541406 Presidente Prudente |
| 6 | 3548708 S. Bernardo do Campo | 3546801 Santa Isabel | 3506003 Bauru | 3506003 Bauru |
| 7 | 3548807 S. Caetano do Sul | 3530607 Mogi das Cruzes | 3541406 Presidente Prudente | 3552205 Sorocaba |
| 8 | 3518800 Guarulhos | 3525904 Jundiaí | 3529005 Marília | 3538709 Piracicaba |
| 9 | 3505708 Barueri | 3552205 Sorocaba | 3555000 Tupã | 3529005 Marília |
| 10 | 3509502 Campinas | 3538709 Piracicaba | 3549904 S. José dos Campos | 3502804 Araçatuba |
| 11 | 3513009 Cotia | 3549904 S. José dos Campos | 3538709 Piracicaba | 3501608 Americana |
| 12 | 3524709 Jaguariúna | 3541406 Presidente Prudente | 3525904 Jundiaí | 3504008 Assis |
| 13 | 3534401 Osasco | 3541000 Praia Grande | 3554102 Taubaté | 3511102 Catanduva |
| 14 | 3549805 S. José do Rio Preto | 3527207 Lorena | 3550605 São Roque | 3524808 Jales |
| 15 | 3549904 S. José dos Campos | 3550605 São Roque | 3546801 Santa Isabel | 3554102 Taubaté |
| 16 | 3552502 Suzano | 3548906 São Carlos | 3540705 Porto Ferreira | 3549904 S. José dos Campos |
| 17 | 3554102 Taubaté | 3554102 Taubaté | 3537602 Peruíbe | 3525904 Jundiaí |
| 18 | 3556453 Vargem Gde. Paulista | 3551009 São Vicente | 3527207 Lorena | 3516200 Franca |
| 19 | 3519071 Hortolândia | 3542602 Registro | 3524303 Jaboticabal | 3524303 Jaboticabal |
| 20 | 3530607 Mogi das Cruzes | 3551702 Sertãozinho | 3518404 Guaratinguetá | 3547809 Santo André |

## DISCUSSION

Regarding Table 2, the three measures capture some cities that do not appear in the first column, namely Fortaleza (CE), Salvador (BA), Campinas (SP), Ribeirão Preto (SP) and Belo Horizonte (MG). They soon had patients with COVID-19, though. Interestingly, the city of Feira de Santana (BA) appears in all columns - it is the second-largest city of the state and connects the capital to the countryside of Bahia^(14)^. Oppositely, the city of João Pessoa, capital of Paraíba state (PB) does not appear in the top 20 of the second column (best measure - see Figure 9), but two other cities from the state do, namely Campina Grande (PB) and Patos (PB). Campina Grande and Patos are among the five richest cities of Paraíba^(15)^. Note that within the context of an epidemic, such cities are potential super spreaders along with the states’ capitals. Five cities of Pernambuco state (PE) appear in the second column (best measure - see Figure 9), namely Caruaru, Carpina, Limoeiro, Paudalho, and Recife. Pernambuco is currently ranked as the second state in the number of confirmed cases of the Northeast region^(4)^.

Table 3, as in Table 2, also displays cities that are captured by the three rightmost columns that do not appear in the first, showing their high level of vulnerability: Ribeirão Preto, Jundiaí, Sorocaba, Piracicaba, and Presidente Prudente. They all have documented cases before June 4th. Our study also captured the most influential cities that had cases already in the beginning, like São Paulo, Campinas, São José do Rio Preto, São José dos Campos and Taubaté. Other cities appear in the second column (best metric) but not in the first: Praia Grande, São Vicente, São Carlos, Registro, Sertãozinho.

Due to their importance in mobility, many cities of Table 3, especially in the second column, appear in the report^(16)^ on the vulnerability of microregions of São Paulo state to the SARS-CoV-2 pandemic of April 5th either as potential spreaders or places with a high probability of receiving new cases. They all have notified cases by June 4th and some have the highest numbers of São Paulo state^(17)^.

Both *s* and *b* with η_0_ pose good results at the beginning of the pandemics for the BRWN network, but *s* alone started to be the best predictor from the end of April. The most important cities, due to their high flow of travelers and their role in the most used routes, are reached first, followed by those with smaller flows, probably because of the interiorization of the virus - the outbreak reaching the countryside cities. This behavior is even more pronounced in SP, in which *s* under η_*d*_ is the best option at first, neglecting lower flow venues, especially in April, but the η_0_ started to be the best option from May onwards.

In the ongoing pandemics, from May 1st, the *s* index with η_0_ is currently the best predictor and may help to figure out which countryside cities are about to receive new cases. Moreover, it may help in the following waves of the disease. In the case of another pandemic, one could first compute the strength of the networks according to the last updated data from IBGE and identify the top-ranked cities. In Brazil, it is enough checking on strength at the original data, as we presented, since it produces similar results as the betweenness centrality and is computationally cheaper to obtain. Regarding the state of São Paulo, one better checks on the strength index with threshold η_*d*_ in the first weeks and only then switch to η_0_. As our results show, the correspondence has statistical significance and, along with other information about the regions such as where are the first notified cases, the pandemic could be closely traced.

## FINAL REMARKS

We present a complex network-based analysis in the Brazilian inter-cities mobility networks towards the identification of cities that are vulnerable to the SARS-CoV-2 spreading. The networks are built with the IBGE terrestrial mobility data from 2016 that have the weekly flow of people between cities. The cities are modeled as nodes and the flows as weighted edges. The geographical graphs, (geo)graphs, are visualized within Geographical Information Systems.

Two scales are investigated, the Brazilian cities without the North region, and the state of São Paulo. The former does not account for the North due to the high number of fluvial routes and some intrinsic local characteristics that are not represented with the terrestrial data. The state of São Paulo is crucial in the ongoing pandemic since the first documented case was in the state capital, and it is currently one of the main focuses of the virus spreading.

Three network measures are studied, namely the strength, degree, and betweenness centrality, under several flow thresholds to account for different mobility intensities, ranging from the original flow data to networks with only the edges with higher weights. We verified that the strength has the best matching to the cities with COVID-19 confirmed cases. Moreover, the strength measure with the original flows showed to be the best option for Brazil. Differently, a more restricted threshold culminates in better correspondences at the beginning of the pandemic in SP. Probably due to the interiorization of the spreading process, a transition is observed after a certain point, around the first days of May 2020, when the original flows have better results as the connections to smaller cities are only present when they are accounted for.

Surprisingly, some countryside cities such as Campina Grande (state of Paraíba), Feira de Santana (state of Bahia), and Caruaru (state of Pernambuco) have higher strengths than some states’ capitals. Furthermore, some cities from the São Paulo state such as Presidente Prudente and Ribeirão Preto are captured in the top-rank positions of all the analyzed network measures under different flow thresholds. Their importance in mobility is crucial and they are potential super spreaders like the states’ capitals.

It is worth mentioning that the COVID-19 data comes from the Brazilian Health Surveillance System, which is fed with data provided by each city in the country. Considering the size and heterogeneity of Brazil, it is important to highlight that there are differences in the capacity to detect cases opportunely and in the quality of the information^(18)^.

As future work, we intend to analyze aerial and fluvial mobility data as well, as they include valuable information about the transport of people and goods. The former is fundamental to the discussion of the dynamics for the Brazilian North region, especially the Amazon, and the latter captures long-range connections. Lastly, one could check for correspondences between the networks’ measures and data from other epidemics, and analyze control measures based on topological properties associated with the mobility network^(19)^.

## Data Availability

The data we used are publicly available and can be found in two publications that are properly cited within the manuscript: 1) IBGE. (2017). Ligacoes rodoviarias e hidroviarias: 2016. Coordenacao de Geografia, Rio de Janeiro, ISBN 9788524044175; 2) W. Cota. Monitoring the number of COVID-19 cases and deaths in Brazil at municipal and federative units level. (2020). Scientific Electronic Library Online (SciELO). DOI https://doi.org/10.1590/SciELOPreprints.362.

## AUTHORS’ CONTRIBUTION

L. S. Freitas and L. B. L. Santos conceived the original idea, methodology, collected the data, interpreted results, wrote the paper and developed the methodology. T. C. R. O. Konstantyner developed the methodology, interpreted results and wrote the paper. J. Feitosa and C. S. N. Sepetauskas developed the methodology. All authors reviewed and approved the final version. The authors declare no conflict of interest.

## ACKNOWLEDGEMENTS

We would like to thank Jussara Angelo (Fiocruz, Rio de Janeiro) for the valuable discussions.

## REFERENCES

1. Li Q, Guan X, Wu P, Wang X, Zhou L, Tong Y, Ren R, Leung KS, Lau EH, Wong JY, et al. Early Transmission Dynamics in Wuhan, China, of Novel Coronavirus–Infected Pneumonia. N Engl J Med. 2020.

2. Worldometers.info. Coronavirus in number. Accessed on May 15th, 2020. [updated 2020 May 15; cited 2020 May 15]. Available from: https://www.worldometers.info/coronavirus/.

3. Coronavírus Brasil. Painel de casos de doença pelo coronavírus 2019 (COVID-19) no Brasil pelo Ministério da Saúde. [updated 2020 May 14; cited 2020 May 14]. Available from: https://covid.saude.gov.br/.

4. Cota W. Monitoring the number of COVID-19 cases and deaths in Brazil at municipal and federative units level. Scientific Electronic Library Online (SciELO). [Internet]. 2020. Available from: https://doi.org/10.1590/SciELOPreprints.362.

5. Lana RM, Gomes MFC, Lima TFM, Honorio NA, Codeço CT. The introduction of dengue follows transportation infrastructure changes in the state of Acre, Brazil: A network-based analysis. PLoS neglected tropical diseases. 2017; 11(11).

6. Angelo JR, Katsuragawa TH, Sabroza PC, Carvalho LAS; Silva LHP, Nobre CA. The role of spatial mobility in malaria transmission in the Brazilian Amazon: the case of Porto Velho municipality, Rondônia, Brazil (2010-2012). PloS one. 2017; 12(2).

7. Saba H, Moret MA, Barreto FR, Araújo Mlv, Jorge EMF, Filho ASN, JGV Miranda. Relevance of transportation to correlations among criticality, physical means of propagation, and distribution of dengue fever cases in the state of Bahia. Sci Total Environ. 2018; 618: 971–976.

8. Santos LBL, Carvalho LM, Seron W, Coelho FC, Macau EE, Quiles MG, Monteiro AM. How do urban mobility (geo) graph’s topological properties fill a map?. Applied Network Science. 2019; 4(1): 1–14.

9. Coelho FC, Lana RM, Cruz OG, Codeço CT, Villela D, Bastos LS, Piontti AP, Davis JT, Vespignani A, Gomes MF. Assessing the potential impact of COVID-19 in Brazil: Mobility, Morbidity and the burden on the Health Care System. medRxiv. (2020).

10. Estrada E. The structure of complex networks: theory and applications, Oxford University Press, (2012).

11. IBGE - Instituto Brasileiro de Geografia e Estatistica, Censo demográfico 2010: Resultados gerais da amostra. [updated 2013 August 08; cited 2020 June 10]. Available from: https://censo2010.ibge.gov.br/resultados.html.

12. Cavararo R. IBGE. Ligações rodoviárias e hidroviárias: 2016. Coordenação de Geografia. Rio de Janeiro: Edgard Blucher; 2017. 79 pp.

13. Ceron W, Santos LB, Neto GD, Quiles MG, Candido OA. Community Detection in Very High-Resolution Meteorological Networks. IEEE Geoscience and Remote Sensing Letters. 2019.

14. Agência IBGE Notícias. IBGE divulga as estimativas da população dos municípios para 2019. [updated 2020 June 14; cited 2020 June 14]. Available from: https://agenciadenoticias.ibge.gov.br/agencia-sala-de-imprensa/2013-agencia-de-noticias/releases/25278-ibge-divulga-as-estimativas-da-populacao-dos-municipios-para-2019.

16. Guimarães Jr PR, Muniz D, Giacobelli L, Maia K, Gaiarsa M, Assis AP, Santana P, Santana EM, Birskis-Barros I, Medeiros L, Velásquez V, Burin G, Marquitti F, Dáttilo W, Cantor M, Lemos-Costa P, Raimundo R, Andreazzi C, Pires M, Côrtes M, Migon E. Vulnerabilidade das microrregiões do Estado de São Paulo à pandemia do novo coronavírus (SARS-CoV-2). Scientific Electronic Library Online (SciELO). [Internet]. 2020. Available from: https://doi.org/10.1590/SciELOPreprints.49.

17. Cidadão - Secretaria da Saúde - Governo do Estado de São Paulo. Novo Coronavírus (COVID-19) Situação Epidemiológica. [Internet]. 2020. Available from: http://www.saude.sp.gov.br/resources/cve-centro-de-vigilancia-epidemiologica/areas-de-vigilancia/doencas-de-transmissao-respiratoria/coronavirus/coronavirus010520_65situacao_epidemiologi ca.pdf.

18. Teixeira MG, Costa MCN, Carmo EH, Oliveira WK, Penna GO. Vigilância em Saúde no SUS - construção, efeitos e perspectivas. Ciência & Saúde Coletiva. DOI https://doi.org/10.1590/1413-81232018236.09032018. 2018; 23(6): 1811–1818.

19. Freitas V, Feitosa J, Sepetauskas CSN, Santos LBL. Robustness analysis in an inter-cities mobility network: modeling municipal, state and federal initiatives as failures and attacks. arXiv preprint 2004.02648. 2020.

